# Weather Shocks, Child Mortality, and Adaptation: Experimental Evidence from Uganda

**DOI:** 10.1101/2023.04.16.23288638

**Authors:** Martina Björkman Nyqvist, Tillmann von Carnap, Andrea Guariso, Jakob Svensson

## Abstract

**Background:** Human-caused climate change is already increasing the intensity and frequency of extreme weather events, such as droughts. The health and economic consequences of these events are expected to be particularly severe for populations in low-income settings whose livelihoods rely on rain-fed agriculture. Within these populations, children are an especially vulnerable group, as undernutrition is linked to 45% of all child deaths across the globe. Despite progress, adaptation gaps exist. We still lack strong evidence on policies to effectively mitigate climate change’s most severe consequences for children. In this paper we ask whether adaptation investments in the form of improved community healthcare can build climate resilience in a low-income country setting.

**Methods:** We reanalyzed data from a clustered randomized controlled trial inducing spatial variation across 214 Ugandan villages in community-health program strengthening, and combined it with quasi-experimental data on locality-specific rainfall shocks. In the intervention clusters, financially incentivized community health workers (CHWs) were deployed over a three-year period to conduct home visits and provide integrated community case management and maternal, newborn, and child health treatment and preventive services. The trial followed 7,018 households with young children (3,790 in 115 intervention clusters and 3,228 in 99 control clusters) over three years. We estimated the effect of low rainfall in the growing season on infant mortality in the following (post-harvest and lean) period, conditional on CHW deployment, over six season-pairs in 2011-2013.

**Findings:** There were 134 infant deaths in the intervention clusters (38.6 deaths per 1000 infant-years) over the three-year trial period. 60 deaths (40.7 deaths per 1000 infant-years) occurred in periods following growing seasons with rainfall below the long-run detrended mean (rainfall deficit seasons), and 74 deaths (36.8 deaths per 1000 infant-years) occurred in periods following growing seasons rainfall above the long-run detrended mean (rainfall surplus seasons). There were 160 infant deaths in the comparison clusters (61.3 deaths per 1000 infant-years). 83 deaths (81.5 deaths per 1000 infant-years) occurred in periods following rainfall deficit seasons, and 77 deaths (46.3 deaths per 1000 infant-years) occurred in periods following rainfall surplus seasons. Adjusting only for the stratified random assignment of clusters, the mean difference corresponded to a 46% reduction in under-five mortality rate (*p*=.000; adjusted rate ratio 0.54, 95% CI 0.39-0.73) following rainfall deficit seasons. The risk of infant deaths in the comparison relative to the intervention group increased in the magnitude of the rainfall deficit.

**Interpretation:** Adaptation investments in a low-income context – here in the form of improved access to community health care – reduced the risk of infant mortality following adverse weather events.

## Introduction

There is a growing consensus that climate change will disproportionately affect countries and populations already facing poverty and malnutrition, and that for these countries and populations in particular, health is a primary channel through which climate change will affect human welfare^*1*^. More frequent extreme weather events, including droughts and heatwaves, are identified as important exposure pathways. Drought poses multiple risks to health. Many of these risks are indirect and affect the circumstances of living through, for example, water shortages and lower crop productivity and income. The nutritional channel – the best recognized health impact of drought – is particularly important in Sub-Saharan Africa, where the majority of the poor rely on rain-fed agriculture for their livelihoods^*2,3*^.

The factors linking drought with increased health risks operate within the context of existing infrastructure and health service provision. Several studies have investigated mortality-mitigating channels, including exploiting quasi-experimental evidence on trade openness^*4*^, credit and liquidity access^*5,6*^, and water and sanitation infrastructure^*7-9*^. Yet, randomized controlled trial-based estimates of effective adaptation policies are missing^10^.

In many developing countries, community health worker (CHW) programs constitute the primary strategy to extend primary healthcare from facilities to underserved rural communities^11^. Systematic reviews of existing studies show that CHW programs can be impactful in promoting positive health behavior and in providing basic curative and health services^12-19^. Two proof-of-principle studies document large reductions in neonatal mortality^20,21^. A study in Uganda where CHWs were financially rewarded for providing integrated community case management (iCCM), maternal, newborn, and child health services (MNCH), and basic treatment and preventive services, reveals similar effects^22^.

In this paper, we combine data from the previously documented trial in Uganda^22^ – inducing spatial variation across localities in community-health program coverage – with data from a natural experiment – inducing variation in growing-season precipitation – to assess whether adaptation investments in the form of community healthcare access can help build climate resilience in a low-income country setting.

### Study Context

In 2007 Living Goods, a US-based NGO active in Uganda, in collaboration with BRAC Uganda, a Bangladesh-based NGO also active in Uganda, began piloting a financially-incentivized community health worker program. Unlike the existing government-run and volunteer-based Ugandan Village Health Team program (VHT), the NGO deployed CHWs and linked monetary rewards to performance tasks such as home visits, preventive health education, provision of basic medical advice, referrals of more severe cases to the closest health center, and sale of subsidized preventive and curative health products. The CHWs were also instructed and financially rewarded to conduct newborn home visits within the first 48 hours of life and to encourage pregnant women to deliver in a facility or with professional assistance.

The study area was predominantly rural, with small-scale, rain-fed, farming being the main source of livelihood. Consumption per adult equivalent in the study districts was US$0.91 per day in 2013^23^. At baseline, community healthcare service was ineffective and in many rural villages de facto non-existent. The Ugandan VHT program received the lowest program functionality score in a WHO review implemented two years before the trial began^12^.

Most of Uganda experiences two rainy seasons associated with the seasonal northward and southward migration of the Inter-Tropical Convergence Zone^23^. The bimodal rainfall pattern gives rise to the country’s bimodal agricultural cycle, with the main rainy seasons (growing seasons) occurring from March to May (MAM) and September to November (SON)^24^. There is considerable spatial and time-series variation with respect to the frequency, and distribution of growing-season rainfall (see supplementary materials (SM), section 1).

### Study design and data

We used two sources of data – a clustered randomized controlled trial (CRCT) and a natural experiment – to assess whether adaptation investments in the form of community healthcare access can help build climate resilience in a low-income country setting. We operationalized resilience as mitigating the adverse impact of rainfall shocks on infant mortality.

The community health program trial was a three-year stratified CRCT initiated in 2011 with a total sample of 214 clusters (rural villages) spread across twelve geographical zones of clusters in ten districts of Uganda. A CHW was assigned to each cluster in the intervention group (115 clusters). No CHW was assigned to the control clusters (99 clusters). Randomization was stratified by geographic zone. The trial’s main objective was to assess the impact on child mortality of having a CHW working in the cluster. A full description of the intervention and trial design is provided in a previously published article^22^. A shorter description of the intervention and trial design is also provided (see SM, section 2).

The primary outcome in the trial was under-five mortality. Given the short exposure windows and the fact that 78% of the child deaths in the comparison group occurred before the child turned one year, we here report results for infant deaths (we provide results for under-five and neonatal mortality rates in SM, section 4.). Infant mortality rates were measured through a cross-sectional household survey administered at the end of the trial, approximately three years after the community health program began operating in the intervention clusters. Mortality rates for each cluster were calculated for six post-harvest exposure periods; i.e., for the six months following the MAM and SON rainy seasons as illustrated in Fig. 1.

**Fig. 1.**
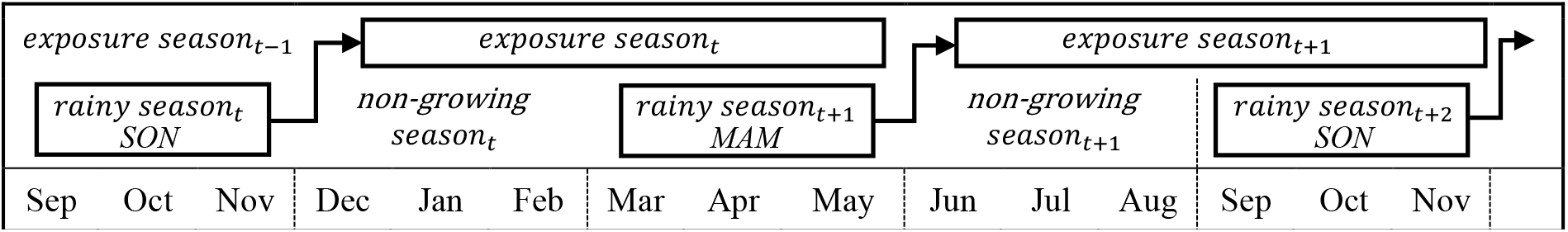
Rainfall and exposure periods. This figure illustrates the study design, linking precipitation in the rainy season *t* to mortality outcomes in the following exposure (non-growing and rainy season) season *t*. The full sample includes data for six rainy seasons (three SON, three MAM) and six exposure seasons.

### Outcomes

A sample household survey at the end of the trial collected birth and death information for all children under five years living in the households as well as for all children under the age of five that had died during the trial period. We used this data to derive cluster-specific infant mortality rates for six exposure periods over the three-year trial period (June-November following the MOM rainy season and December-May following the SON rainy season). The infant mortality rate was calculated as the number of deaths within the first year of life during each exposure period per 1000 infant-years of exposure, with infant-months of exposure defined as the difference between the birth date of the child, or the start date of the exposure period, if the child was born before that date, and the date that the child turned one year, if that occurred during the exposure period, or the date of the end of the exposure period if the child was less than one year, or the date of the death of the child. The same approach was used to estimate under-5 mortality. For neonates, mortality was expressed per 1000 births.

We used the TAMSAT dataset to derive the rainfall measures, since rain gauge-based data with fine temporal and spatial scales were lacking^26^. The TAMSAT data – with rainfall estimates based on satellite data measuring changes in cloud surface temperature to infer discharged volumes – is unique in its detailed spatial (0.0375 × 0.0375 degree earth grid or 4.2 × 4.2 kilometers at the equator) and temporal (daily estimates) resolution^27^. Validation exercises have found satisfactory accuracies of these predictions relative to ground measures^24^. We derived a localized rainfall index by matching trial villages with the rainfall profile of the grid cell they fell into. For each season type *s, s* = {*MAM, SON*}, we calculated the cumulative amount of precipitation 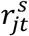 per cluster (*j*), season type (*s*), and year (*t*), over a 20-year period (December 1993 - November 2013). As the data displayed a secular trend towards more rainfall over time, we used a linear regression model to detrend the rainfall data and denoted the detrended rainfall as 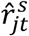. Finally, we calculated the standard deviation of the detrended time series (denoted 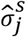) and define the rainfall index (*RI*) as 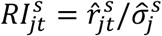; i.e., the normalized deviation from the long-run detrended mean in local rainy season precipitation. The rainfall index is interpreted as quasi-random with regard to cluster-level outcomes and is similar to the Standardized Precipitation Index (SPI), a widely-used measure to define (agricultural) drought conditions^28^, but based on the cumulative rain during the growing season rather than on monthly rainfall. We also used the rainfall index to define rainfall deficit and rainfall surplus seasons, denoted 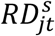 and 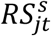, where 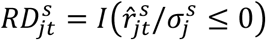, with *I*(.) an indicator function that is 1 if its argument is true and 0 otherwise. That is, a rainfall deficit season was a season with rainfall below the long-run detrended mean, and a rainfall surplus season, 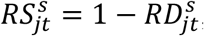, was a season with rainfall above the long-run detrended mean.

### Statistical analysis

We linked rainfall in rainy season *t* to infant mortality in the following six-month exposure season *t* over a three-year (six seasons) period, as illustrated in Fig. 1. For our cluster-level analyses, this procedure results in an effective sample size of 6 seasons x 214 clusters = 1,284 cluster-season observations. We report intention-to-treat estimates, with the intention to treat being defined by the cluster of residence at baseline. We used three specifications (see SM section 3). First, infant mortality rates conditional on assignment status (treatment and control groups) and rainfall status (rainfall deficit or surplus season) were reported, with rate ratios computed using a Poisson model, adjusting for the stratified randomized design using binary geographic zone indicators. Second, we used a linear interaction specification with cluster and exposure season-specific infant mortality modeled as a function of the assignment status, the rainfall index (*RI*), and their interaction. Finally, exploiting the same interaction specification, we estimated the risk of infant death using a logit model, controlling non-parametrically for exposure length using age-bins indicators, and derived relative risk ratios conditional on the magnitude of the rainfall shock. In all specifications, we reported unadjusted estimates, i.e., we only accounted for the stratified random design. Standard errors were clustered at the rainfall grid cell level to account for intra-cluster correlation across households exposed to the same rainfall shock.

## Results

The analysis was based on a sample of 7,018 households that have lived in the same cluster throughout the trial, and their 7,899 children aged less than one year at some point during the trial period. The mean number of households per cluster was 237 at baseline. Baseline households and cluster characteristics, and in- and out-migration rates, were evenly distributed between the two assignment groups as previously described^24^. Summary statistics for the rainfall index and the binary rainfall deficit and surplus seasons are provided (see SM section 1).

In Table 1 we begin by splitting the sample into exposure seasons following a rainfall deficit and a rainfall surplus season. In the control group, the infant mortality rate following a rainfall deficit season was 81.5, compared to 46.3 following a rainfall surplus season. In the treatment group – with CHWs rewarded for delivering timely and appropriate basic health services – mortality rates following a rainfall deficit and rainfall surplus season were broadly similar: 40.7 compared to 38.6. Adjusting only for the stratified random assignment of clusters, the mean difference corresponded to a 46% reduction in infant mortality following a rainfall deficit season – an effect significant at the 1% level. While mortality rates were lower in the intervention relative to the control group also following a rainfall surplus season, we cannot reject the null hypothesis that mortality rates were the same in the two groups. We document similar results for under-5 and neonatal mortality (see SM, section 4)

**Table 1.** Infant mortality: summary statistics and rates conditional on assignment status and rainfall outcomes. All data are from the endline sample household survey. Table 1 displays total years of exposure to the risk of death for infants and deaths under 1 year in the sample, and mean infant mortality rate (number of infants deaths per 1,000 years of exposure to risk of death for infants) across cluster-by-exposure seasons. The adjusted rate ratios; i.e., the infant mortality rate in the treatment relative to the control group, adjusting for the stratified randomized design using binary geographical zone indicators, conditional on rainfall status, are computed using a Poisson model. Standard errors, in parentheses, are clustered at the rainfall grid cell level. Significance: *\*P <* 0.10, *\*\*P <* 0.05, *\*\*\*P <* 0.01.

| Rainfall deficit season (*RD*)<br>Assignment group | RD = 1 |  | RD = 0 |  |
| --- | --- | --- | --- | --- |
|  | Control | Treatment | Control | Treatment |
| Years of exposure to risk of death | 1197 | 1510 | 1808 | 2033 |
| Deaths under 1 year | 83 | 60 | 77 | 74 |
| Mortality rate per 1000 years of exposure | 81.5 | 40.7 | 46.3 | 36.8 |
| Adjusted rate ratio | 0.538***<br>(.085) |  | 0.809<br>(.131) |  |
| Observations | 1284 |  | 1284 |  |

The differential impact of rainfall shocks across assignment groups is illustrated in Fig. 2 and further documented using an interaction model in Table 2, column (1). Again, similar patterns are observed for neonatal and under-5 mortality (see SM section 4).

**Table 2:** Infant mortality and risk of infant death conditional on rainfall and assignment status. Column (1) displays estimates from a linear regression where the cluster-by-season infant mortality rate is regressed on the rainfall index, the assignment status, and their interaction. Column (2) displays estimates from a logit model where the binary outcome is infant death, and where exposure length is controlled for non-parametrically using 12 binary age-bin indicators. Binary geographical zone indicators are included in both specifications to adjust for the stratified randomized design. Standard errors, in parentheses, are clustered at the rainfall grid cell level. Significance: *\*P <* 0.10, *\*\*P <* 0.05, *\*\*\*P <* 0.01.

|  | Infant mortality |  |
| --- | --- | --- |
|  | (1) | (2) |
| Rainfall index (*RI*) | -14.02**<br>(5.65) | 0.793**<br>(.094) |
| Treatment group | -29.92***<br>(7.44) | 0.614***<br>(.086) |
| *RI*×Treatment group | 16.54***<br>(6.10) | 1.33**<br>(.176) |
| Observations | 1284 | 14,876 |

**Fig. 2.**
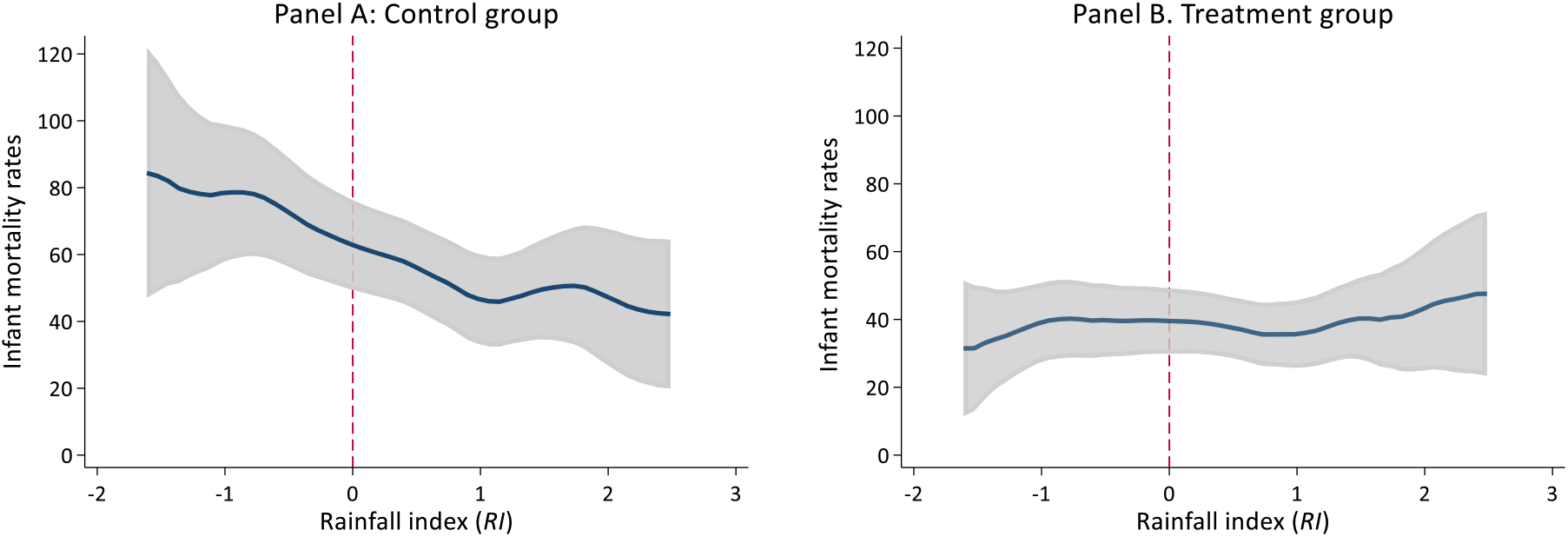
Infant mortality and rainfall shocks conditional on assignment status. Smoothed values from a local polynomial regression of infant mortality (six months-windows) on deviations in rainfall from the long-run detrended mean (three months-windows) in control and treatment groups. Gray-shaded areas represent 95-percent confidence intervals.

To further investigate the relationship between negative rainfall shocks and infant mortality, and the extent to which adaptation investments in the form of community-health strengthening could have mitigated these health shocks, we estimated the relationship between the risk of infant death and the magnitude of the rainfall shocks (Table 2, column (2)), and evaluated the risk at different values of the rainfall index (Fig. 3), with results for neonatal and under-5 mortality reported in SM, section 4.

**Fig. 3.**
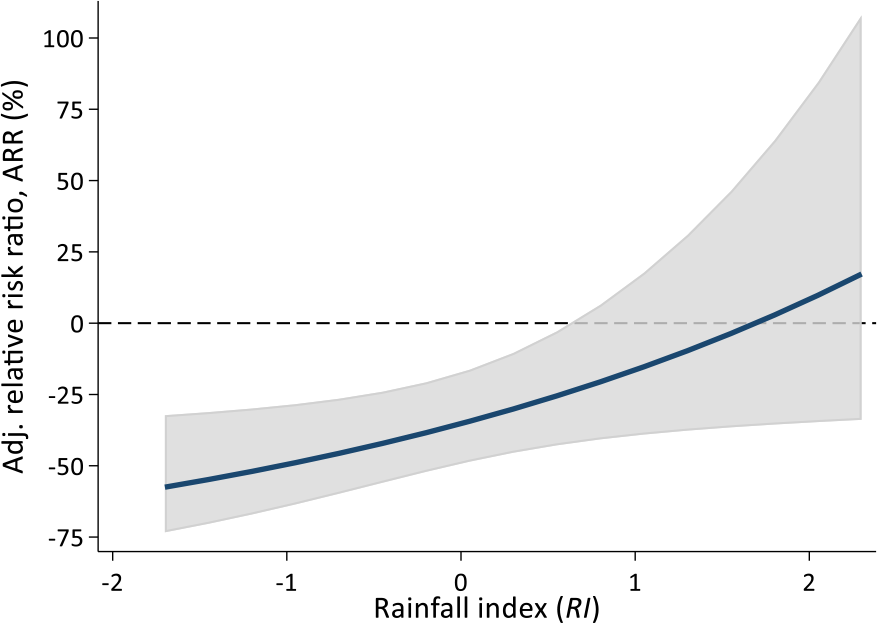
Relative risk of neonatal death conditional on rainfall shocks. This figure displays adjusted risk ratios from an interaction model (see Table 2, column 2) for different values of the rainfall index Gray shaded area represents 95 percent confidence intervals, with standard errors clustered at the rainfall grid cell level.

The relative risk of infant deaths between the treatment and control groups was increasing in the absolute magnitude of a negative rainfall shock. At the threshold for a moderate drought (*RI* = −0.8), following the classification in the U.S. Drought Monitor^29^, infants in the treatment group were 47% less likely to die than infants in the control group (adjusted risk ratio 0.53, 95% CI 0.39-0.73).

### Interpretation and discussion

Climate change is expected to worsen the frequency, intensity, and impacts of extreme weather events. The impact is predicted to disproportionately affect countries and populations already facing poverty, malnutrition, and poor health service provision. In this paper, we provided evidence consistent with this prediction: the risk of infant death increased following adverse growing-season rainfall shocks. We also showed that adaptation investments in the form of strengthened community-health service provision could significantly dampen these risks.

The results should be interpreted within the context of the study. First, the intervention was implemented across villages lacking access to effective community-health service provision at the onset of the study, and the quality of the public primary healthcare system was notoriously low^30^. Furthermore, the sample population was poor. These characteristics could help explain the large magnitudes of the effects we document, but also highlight the inherent potential as well as feasibility to support climate resilience by strengthening community healthcare provision. Second, we observed few extreme events in the data. A SPI ≤ -0.8 is commonly used as a threshold for a moderate drought, following the classification in U.S. Drought Monitor, with lower thresholds for severe, extreme, and exceptional droughts. In the data we observed no cluster-season episode classified as an exceptional drought (i.e., RI < -2), while 1.5% of the episodes were classified as an extreme drought (−1.9 ≤ RI ≤ -1.6). Thus, predictions about the mitigating impact of community-health program strengthening in the case of the most extreme weather events are uncertain. The results, however, also suggest that for a poor rural population, less extreme deviations in growing season rainfall significantly raise the risk of infant death and improved community-health services provision can play an important role in reducing that risk. By design, we focused on the reduced form effect of a rainfall shock. Thus, we did not differentiate between rainfall-induced shocks occurring in utero – through, for example, maternal malnutrition in the second and third trimester – or during early childhood. We also note that low rainfall affects the circumstances of living in several ways, including through threats to drinking water supplies, sanitation, spread of vector-borne and water-borne diseases, and impacts on livelihoods. These circumstances could have led to a series of health effects resulting in increased morbidity and mortality. Lower crop output and/or loss of livestock may have also changed the tradeoff within the household between infant care and income-generating activities, which in turn could have exacerbated the effect of an adverse weather-induced shock on infant health. Weather shocks could also have resulted in prices of outputs and inputs changing, dampening the income fall of an adverse shock to agricultural output. The focus on local rainfall shocks also implied that locally-based informal institutions for risk-pooling and insurance – which could have played an important role in mitigating idiosyncratic shocks to income – were likely not effective. Thus, our core focus was not on specific mechanisms through which low rains and drought conditions affected child mortality, but rather on whether, and to what extent, improved access to iCCM, MNCH, and basic preventive and treatment services mitigated the adverse consequences of extreme weather events.

Time-series data on intermediate health service outcomes were not available. Based on household survey data collected at the end of the three-year trial period, however, the community-health service intervention resulted in large increases in both the quantity and, more suggestively, the quality of several maternal, newborn, and child health services^22^.

In this paper, we provided to our knowledge the first causal evidence based on a randomized controlled trial that improved access to basic primary care could play an important role in mitigating some of the adverse predicted consequences of climate change in a low-income setting. Our results provided confirmation that adaptation investments – here in the form of improved access to community care – could protect the most vulnerable from adverse weather events.

## Data Availability

All data produced in the present study are available upon reasonable request to the authors

https://www.dropbox.com/s/9fc8w4sp2r33hi0/Supplementary%20material_medrxiv.pdf?dl=0

## Acknowledgments

We thank Seema Jayachandran, Tessa Bold, and several seminar participants for helpful comments.

## Funding

Children Investment Fund Foundation (MBN, AG, JS)

Riksbankens Jubileumsfond M16:0345:1 (JS)

Handelsbankens Forskningsstiftelser P20-0242 (TvC, JS)

Mistra, the Swedish Foundation for Strategic Environmental Research (MBN)

## Author contributions

Conceptualization: MBN, AG, JS

Methodology: AG, TvC, JS

Investigation: AG, TvC, JS

Visualization: AG, TvC, JS

Funding acquisition: MBN, AG, JS

Project administration: MBN, AG, TvC, JS

Supervision: MBN, AG, TvC, JS

Writing – original draft: JS

Writing – review & editing: MBN, AG, TvC, JS

## Data and materials availability

All data, code, and materials used in the analysis will be made available upon publication as supplementary material, as well as through the personal webpages of the researchers. The views expressed in this article are not necessarily those of the partner organizations, funders, or their members. All errors are our own.

## Supplementary Materials

Click here

